# Molecular Epidemiology and Histopathological Classifications of Lung Cancer in Africa: A Scoping Review Protocol

**DOI:** 10.1101/2022.06.25.22276891

**Authors:** Emmanuel Akomanin Asiamah, Mbuzeleni Hlongwa, Kennedy Nyamande, Themba Geoffrey Ginindza

**Affiliations:** Discipline of Public Health Medicine, School of Nursing and Public Health, University of KwaZulu-Natal, Durban, 4001, South Africa; Cancer and Infectious Diseases Epidemiology Research Unit (CIDERU), College of Health Sciences, University of KwaZulu-Natal, Durban, 4001, South Africa; Department of Pulmonology, School of Medicine, University of KwaZulu-Natal, Durban, 4001, South Africa; Department of Medical Laboratory Sciences, School of Allied Health Sciences, University of Health and Allied Sciences, PMB 31, Ho, Ghana; Burden of Disease Research Unit, South African Medical Research Council, Cape Town, South Africa

**Keywords:** Lung cancer epidemiology, histopathological classifications, early detection, molecular biomarkers, Africa

## Abstract

**Introduction:** Lung cancer is a leading cause of death globally and an emerging epidemic in Africa. Recent advances in molecular biomarkers and understanding of the cancer epidemiology and population-based genomic profile for early detection, diagnosis and treatment show promise in reducing incidence and mortality rate. However, this is lacking in Africa. The main objective of this scoping review is to map the evidence on lung cancer molecular epidemiology, genomic profile, and histopathological distributions in Africa.

**Methods and analysis:** This review will be guided by Arksey and O’Malley’s framework and Levac et al.’s recommendation for methodological enhancement for scoping review studies. A search for keywords from scientific databases (PubMed/MEDLINE, EBSCOhost, SCOPUS and Google Scholar) and grey literature will be conducted for evidence on the molecular epidemiology and histopathological classifications of lung cancer in Africa. The Preferred Reporting Items for Systematic reviews and Meta-Analyses (PRISMA)-Scoping Review Extension guidelines will be used to report screened results. We will use the PRISMA-ScR checklist to ensure the study adheres to sound methodological rigour acceptable for scoping reviews. The study’s search strategy will include Boolean terms (‘AND’ and ‘OR’) and Medical Subject Heading (MeSH) terms.

**Ethics and dissemination:** This review will not include animal or human participants. Ethics approval and consent to participate are not applicable. Findings of this scoping review will be disseminated via electronic/social media, conferences, meetings with stakeholders and peer-review publications.

**Strengths and limitations of the study:**

▸ This review will be the first to identify and map evidence that assesses the molecular epidemiological diversity and histopathological distributions of lung cancer in Africa.
▸ The summary of evidence obtained from relevant studies in the African context will help understand the genetic diversity of lung cancer among high-risk heterogenous African populations and guide future research towards early detection and targeted therapy in the African context.
▸ The review will include all available studies in the literature with no time or language restrictions. Thus, studies published in languages apart from English will be translated using an online tool.
▸ There will be no quality appraisal of the included studies.

## Background

Lung cancer (LC) remains the highest cause of cancer-related deaths worldwide [1]. According to recent reports by the World Health Organization (WHO), lung cancer morbidity and mortality rates were estimated at 2.2 million new cancer cases and 1.8 million deaths, representing approximately 1 in 10 (11.4%) cancers diagnosed and 1 in 5 (18.0%) deaths [2, 3]. The current WHO classification categorises LC into two main groups: small cell lung cancer (SCLC) (15%) and non-small cell lung cancer (NSCLC) (85%), which is further divided into adenocarcinoma (ADC), large cell carcinoma (LCC) and squamous cell carcinoma (SCC) [4-6].

Tobacco smoking is the leading modifiable risk factor for LC [7]. Recent studies have shown that secondary exposure to tobacco products and other environmental factors, including indoor and outdoor air pollutants such as radon and asbestos, are important carcinogens [8]. The incidence and mortality rates of LC are estimated to be three to four-fold higher among populations with increased tobacco consumption [9]. This trend has been noted in high-income countries (HICs) – such as the USA, where it was reported to be responsible for 1 in 5 deaths [10] compared to low-and-middle-income countries (LMICs), including Africa, in the past four decades. Tobacco use in HICs has been declining steadily while rising and becoming an epidemic in Africa [11, 12]. Given that 80% of smokers aged ≥ 15 years currently reside in LMICs [13, 14], this paints a gloomy picture; a projection of 11% of all deaths attributable to tobacco-related diseases in developing countries by 2025 [15]

Also, studies have shown that LC incidence and mortality rates are inversely related to the survival rate [16-18]. A significant setback to the successful treatment of lung cancer for optimal health is attributed to the late presentation of the disease, usually at the advanced stage (stage IIIB/IV), with some evidence of spread, at the time of diagnosis [19]. Thus, impacting clinical outcomes with improved patients’ survival will primarily be driven by the availability of cost-effective, non-invasive screening techniques for efficient and rapid diagnosis. However, these interventions are lacking among African populations [20].

Several radiological screening techniques, including chest x-ray, computed tomography (CT) scan of the chest and thorax, bronchoscopy, ultrasound of the chest and lungs, magnetic resonance imaging (MRI), and positron emission tomography (PET), have been used to aid the diagnosis of lung cancer in HIC countries, through the detection of suspicious lesions and sampling of mediastinal lymph nodes [21-25]. Lung biopsies are obtained by bronchoscopy and surgical procedures for histopathological analysis and appraisal. However, radiation exposure and tissue invasion risks associated with these techniques increase concerns [26, 27].

Several studies have identified a list of biomarkers implicated in lung carcinogenesis, the utility of which has allowed for early detection and distinguishing variants and subtypes of lung cancer [6, 28]. A study by Kang and colleagues reported the β-chain of human serum HP (haptoglobin) as a reliable biomarker for the definitive diagnosis of LC [29]. Other studies have also investigated biomolecular markers and their associated gene mutations, including EGFR, ALK, ROS1, MET and PD-L1, which have been identified to possess prognostic and therapeutic values for LC [30-32]. Current evidence has also shown the diagnostic potential of using biomarkers for the early detection of cancers. A study by researchers in the US showed an overall decrease in death from LC. Among men, the report revealed a decrease of 3.2% from 2008 to 2013 and 6.3% annually from 2013 to 2016 [33].

These clinical and diagnostic innovations are underpinned by the understanding of the molecular epidemiology as well as a robust database on the genomic profile of LC generated by studies conducted mainly in HICs showing the potential use of readily available and easily accessible bodily fluids (including sputum, saliva, urine, and blood) [34-37] at the initial stages of the LC. Consequently, these studies have provided vital information for designing and developing cost-effective, non-invasive diagnostic tools and effective targeted therapy for reducing LC mortality [38, 39]. These developments will contribute to efforts towards achieving the sustainable development goals (SDGs) 3, by the year 2030, with a target of reducing by one-third early mortality from non-communicable diseases (NCDs) through prevention, early detection and treatment and promoting overall quality health and wellbeing [40]

Conversely, there is a paucity of knowledge on the extent of information on molecular epidemiology, ethnic diversity, and tumour heterogeneity in terms of histopathological classifications of LC in Africa. This evidence is vital for developing interventions for early detection and the design of therapeutic target algorithms. To this end, a scoping review of the literature will be conducted to establish the evidence of the molecular mechanisms of genetic diversity and tumour heterogeneity of lung cancer among African populations. This scoping review aims at mapping evidence on the molecular epidemiology and histopathological classifications of lung cancer in Africa.

## Methods

### Protocol design

As part of doctoral research investigating the utility of molecular biomarkers for early detection of lung cancer and genomic profiles among African populations, we will conduct a scoping review of published peer-reviewed and grey literature (comprising theses/dissertations, technical reports, and other articles not indexed in traditional databases, including the WHO library and Google search engine). The proposed review, slated for July to September 2022, will rely upon Arksey and O’Malley’s framework [41] and Levac et al.’s methodological enhancement for scoping reviews [42]. The following Arksey and O’Malley’s scoping review framework stages will guide this review: stage 1: identifying the research question; stage 2: identifying relevant studies; stage 3: eligibility criteria and study selection; stage 4: charting the data; stage 5: collating, summarising and reporting the results.

### Stage 1: Identifying the research question

The Population/Concept/Content (PCC) framework recommended by the Joana Briggs Institute (JBI) [43] will define the eligibility criteria of the studies for the main research question, as shown in Table 1.

**Table 1:**
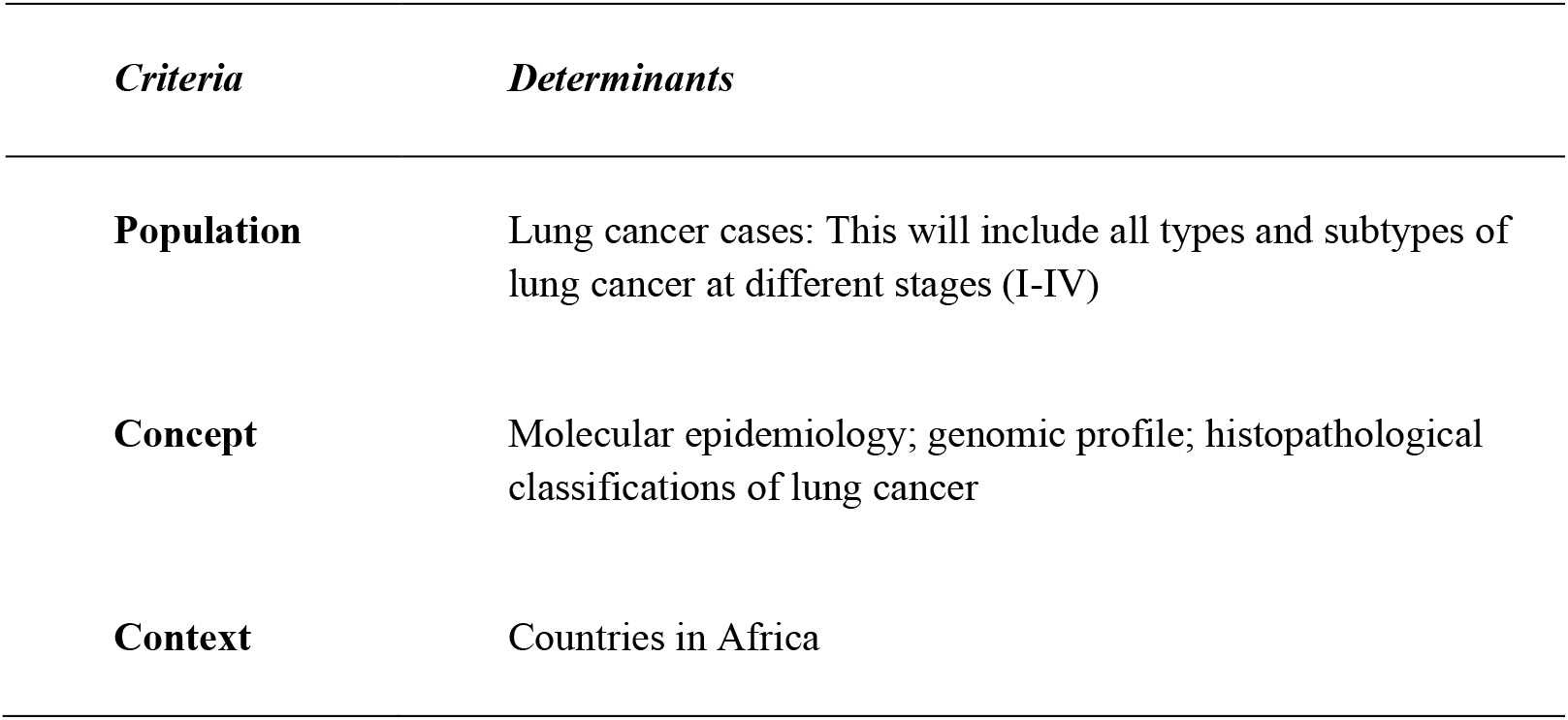
PCC framework for determining research question eligibility.

| *Criteria* | *Determinants* |
| --- | --- |
| **Population** | Lung cancer cases: This will include all types and subtypes of lung cancer at different stages (I-IV) |
| **Concept** | Molecular epidemiology; genomic profile; histopathological classifications of lung cancer |
| **Context** | Countries in Africa |

#### Thus, the main research question for the scoping review is

What is the current scientific evidence and understanding of the epidemiology and molecular mechanism, host factors and genomic profile of lung cancer among African populations?

#### The sub-review questions will be as follows

- What is the current scientific evidence on lung cancer epidemiology and molecular mechanism, host factors, and genomic profile in Africa?
- What is the spectrum of histopathological classifications of lung cancer in Africa?

#### Proposed hypothesis for the scoping review

Evidence of the molecular epidemiological profile and histopathological patterns of lung cancer is lacking in Africa.

### Stage 2: Identifying relevant studies

The search strategy recommended by JBI will be followed to identify relevant studies. A comprehensive search for a primary source of literature on multiple electronic databases, including PubMed/MEDLINE, EBSCOhost, SCOPUS and Google Scholar, will be conducted. A range of search terms and keywords, including “molecular epidemiology”, “genomic profile”, “histopathological classifications”, “lung cancer”, and “Africa”, will be applied in all the searches from the electronic databases. Boolean terms ‘AND’ and ‘OR’ will separate the keywords. Medical Subject Heading (MeSH) terms will also be included in the search string to identify all relevant studies. A further search for additional relevant studies from the reference list of the included articles will be performed and assigned for screening. We will also search for potentially relevant information (grey literature).

An initial pilot search was performed to assess the feasibility of the proposed review; we employed a draft search strategy for PubMed/MEDLINE, as depicted in Supplementary Table 1. (Appendix)

### Stage 3: Eligibility criteria and study selection

Eligibility for the scoping review will be defined by the inclusion and exclusion criteria as follows:

#### Inclusion criteria

The inclusion criteria will encompass:

- Studies on lung cancer conducted among African populations
- Studies presenting evidence on the molecular epidemiological profile of lung cancer
- Studies presenting evidence on histopathology classifications of lung cancer
- Studies presenting evidence on biomarkers for lung cancer
- Studies published in all languages
- Studies based on quantitative and mixed-methods study designs

#### Exclusion criteria

- Lung cancer studies not from the African continent
- Commentaries, editorials, reviews, abstracts, and others.
- Studies conducted by qualitative method study design^1^

#### Study Selection

A three-phase screening approach will capture relevant studies in the proposed scoping review. The principal investigator (PI) will perform the database search, conduct title screening and export selected studies into the EndNote version 20 library. Duplicate articles found in the reference manager will be deleted. The PI and a co-screener will independently conduct abstract screening, followed by full-text screening using pretested google forms. Any disagreements between the two independent reviewers will be resolved through discussion or by a third reviewer when the paired reviewers cannot resolve their conflicts. For articles we could not find in full text, we will contact the University of KwaZulu-Natal’s library services for expert and technical support, including emails to the corresponding authors to access articles we could not retrieve in full text. PRISMA 2020 flow diagram for systematic reviews that include searches of databases, registers, and other sources [44] will be used to account for the articles in each screening phase, as shown in Appendix (Supplementary Figure1).

### Stage 4: Charting the data

A data charting form will be developed and pretested for this scoping review to ascertain its comprehensiveness in extracting all relevant data from the included studies. This form will contain the following information: author(s), publication year, study title, objective/aim; study design; country; study setting, and recommendations/limitations, as shown in Appendix (Supplementary Table 2).

Two independent reviewers will conduct the data extraction from the included studies, and if there are discrepancies to be addressed, a third reviewer will be engaged to resolve them independently. We will use Cohen’s Kappa statistics to assess inter-rater reliability to measure the degree of agreement on selected articles [45]. As the scoping review process evolves, the data extraction form will be modified and updated periodically with reviewers’ feedback.

### Stage 5: Collating, summarising and reporting the results

After charting the data, a descriptive overview of the studies will be presented in a narrative format. An account of the number of studies, research methods used, gender percentage, epidemiological profile and histopathological distributions of lung cancers and clinical outcomes will be described in this narrative. We will map the country-specific outcomes emerging from the studies for the analysis, and a numerical summary of these outcomes will be conducted. Also, a table will be produced to show the numerical summary of the outcomes. A comparison of lung cancer incidence and mortality amongst the African countries will be conducted to analyse the outcomes. The results section will show a geographical map of the individual countries reported in the studies and charts showing the types and subtypes of lung cancer and their available genomic profiles. We will also use NVivo version 12 to organise the themes and sub-themes [46, 47]. The proposed review will be reported according to the PRISMA extension for scoping reviews (PRISMA-ScR) checklist and explanation [48] as shown in Appendix (Supplementary Table 3).

### Ethics and dissemination

The proposed scoping review will not include animal or human participants. Ethics approval and consent to participate are not applicable. All findings will be disseminated via electronic media, conferences, and meetings with stakeholders and published in peer-reviewed journals.

### Patient and public involvement

There were no patients and public involvement in this protocol.

## Discussion

The burden of lung cancer is increasing worldwide. However, recent studies conducted on HICs in the East and West regions of the globe have predicted a significant decline in incidence and mortality rates [34-37]. Proposed models for predicting an optimistic future epidemiological profile of lung cancer through 2030 are currently being explored in clinical trials [49, 50]. Furthermore, current developments in tobacco control programs for LC prevention [14] have shown the prospects of reducing LC mortality in the high-income counties (HICs), as opposed to recent reports of increasing LC burden in Africa [20], which can be attributed to the non-existence of such pragmatic interventions. Also, the potential utility of novel biomarkers isolated from liquid biopsy (including blood, saliva, urine, and sputum) for early detection of LC and identifying driver mutations for targeted therapy has been widely reported in recent studies in HICs [34-36]. On the contrary, this is lacking in most African settings [20]. The proposed scoping review will consolidate evidence on the histopathological subtypes of lung cancer, genomic profiles and associated biomarkers, and their expression patterns among African populations as reported in the literature.

Limitations to this study may include a potential miss of relevant articles due to the exclusion of qualitative studies that do not align with the study design and review articles with relevant studies but cannot be considered for this study.

Ultimately, the results of this scoping review (which is a part of the main study in KwaZulu-Natal, South Africa) anticipate identifying literature gaps, which may inform the methodology of future research for the understanding of the molecular mechanisms and host factors and the histopathological distributions of lung cancer in Africa. This will provide critical information for developing tools for early diagnosis and designing population-based genomic profiles for targeted therapy in Africa. Thus, the study outcome will strengthen the strategies put in place by the Multinational Lung Cancer Control Program (MLCCP) in UKZN for curbing lung cancer incidence and mortality, thereby consolidating efforts toward establishing national policy for annual lung cancer screening among high-risk groups. In addition, the generation of baseline data with the molecular profiling of lung cancers will catalyse targeted therapy development for precision medicine among African populations.

## Data Availability

All data generated or analysed during this study will be made available, including citations to all studies with data that has been presented in the form of references shall be made available upon reasonable request to the authors.

## Abbreviations

ADC: Adenocarcinoma
ALK: Anaplastic Lymphoma Kinase
CT: Computed Tomography
EGFR: Epidermal Growth Factor Receptor
HICs: High-Income Countries
LC: Lung Cancer
LCC: Large Cell Carcinoma
LMICs: Low-and Middle-Income Countries
MRI: Magnetic Resonance Imaging
NCDs: Non-Communicable Diseases
NSCLC: Non-Small Cell Lung Cancer
PCC: Population, Content, and Context
PET: Positron Emission Tomography
PRISMA: Preferred Reporting Item for Systematic Reviews and Meta-Analysis extension for Scoping Reviews
SDGs: Sustainable Development Goals
UKZN: KwaZulu-Natal
WHO: World Health Organisation

## Acknowledgements

The authors would like to thank the Cancer Infectious Diseases Epidemiology Research Unit (CIDERU) for the training and technical support and the University of KwaZulu-Natal for providing resources for this scoping review protocol.

## Authors’ Contributions

EAA, KN and TGG conceptualised the study. EAA, KN and TGG designed the study methodology. EAA wrote the protocol and final draft manuscript. EAA, MH, KN and TGG reviewed the protocol and final draft manuscript. All the authors approved it.

## Data availability

All data produced in this study shall be made available upon reasonable request to the authors.

## Funding statement

This research received no specific grant from any funding agency in public, commercial or not-for-profit sectors.

## Competing interests statement

None declared

## Patient consent for publication

Not required

## APPENDIX: Supplementary Files

**Supplementary Table 1:**
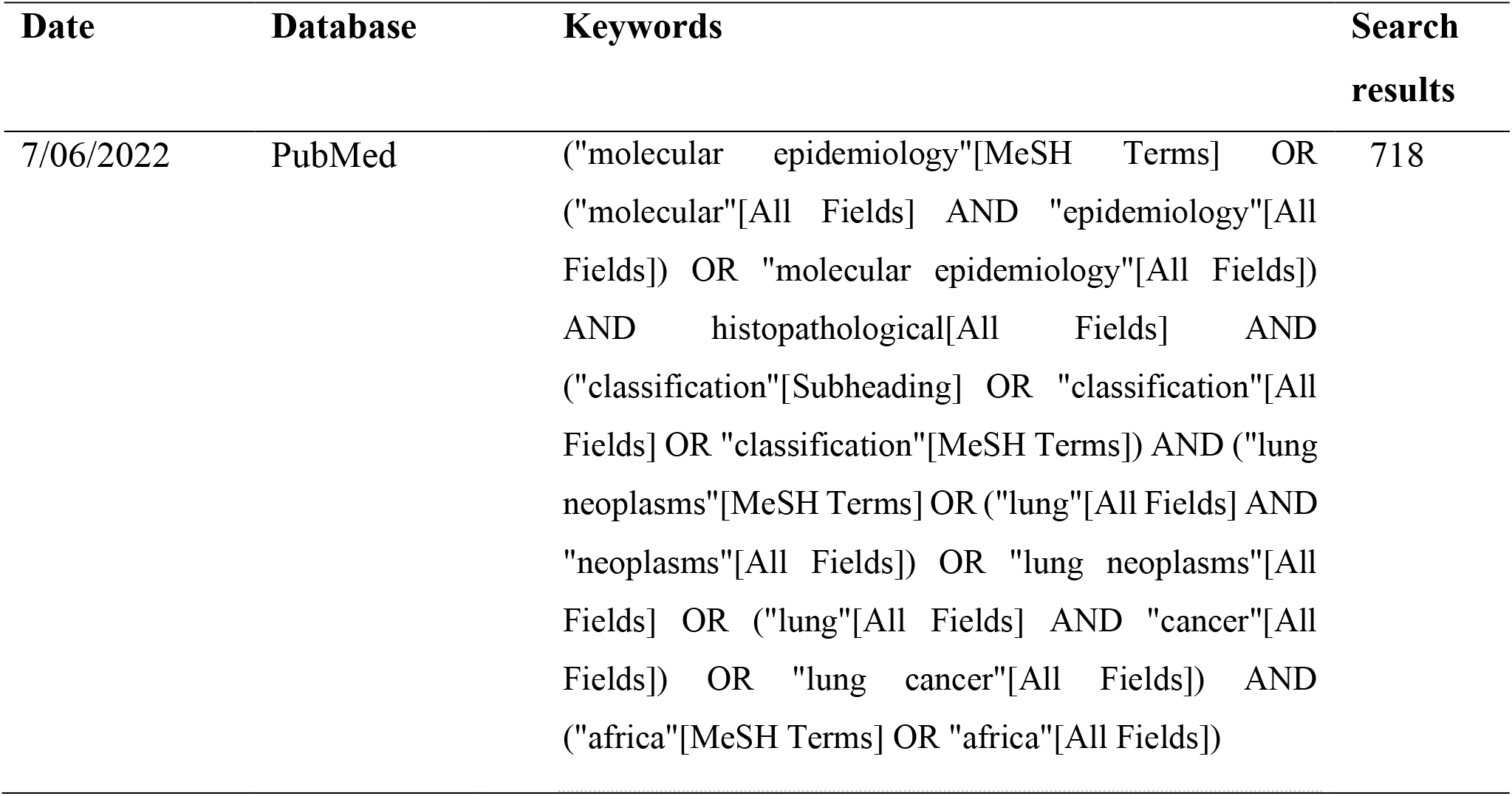
Pilot search in PubMed electronic database.

**Supplementary Table 2:**
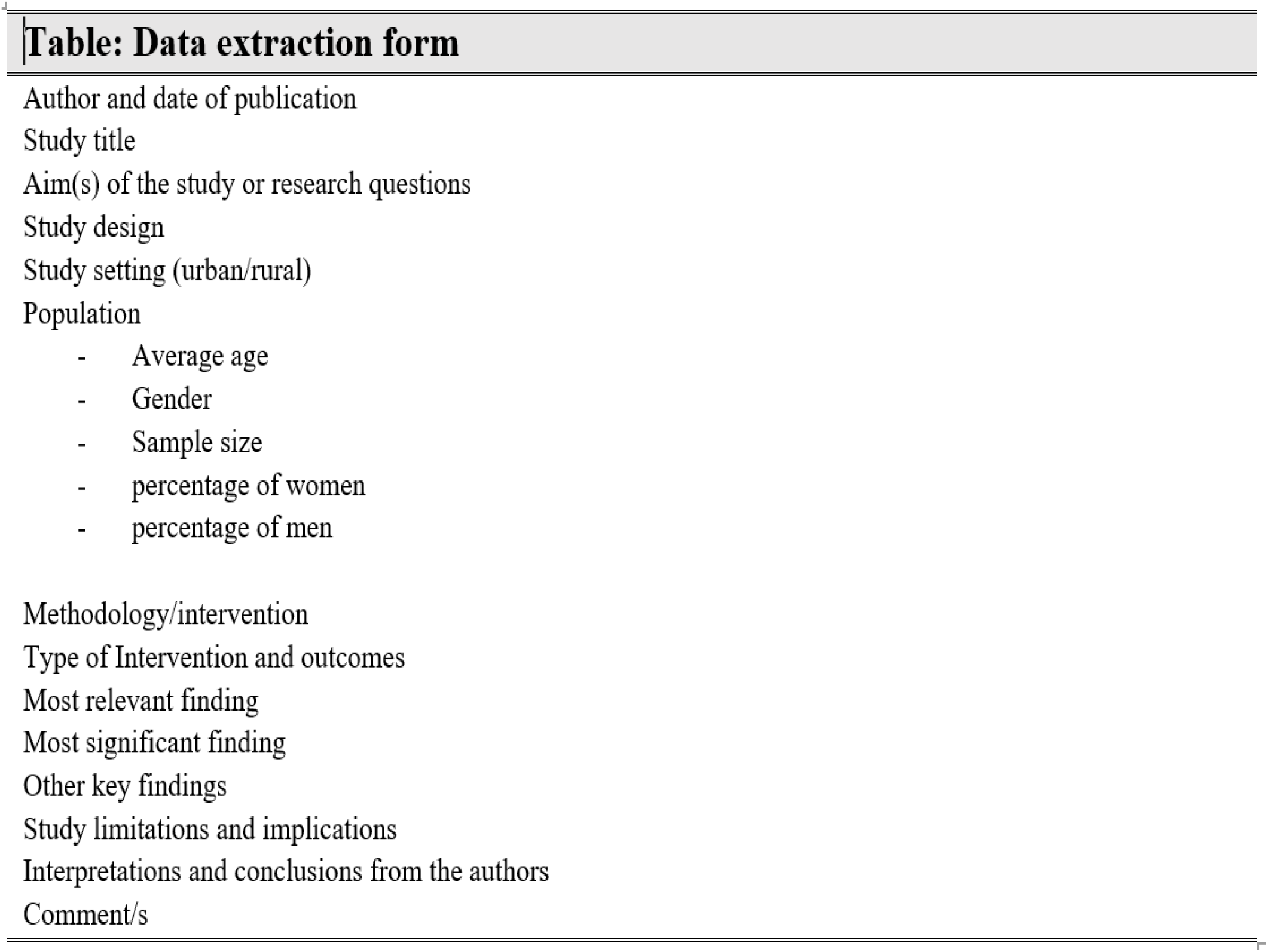

**Supplementary Table 3:**
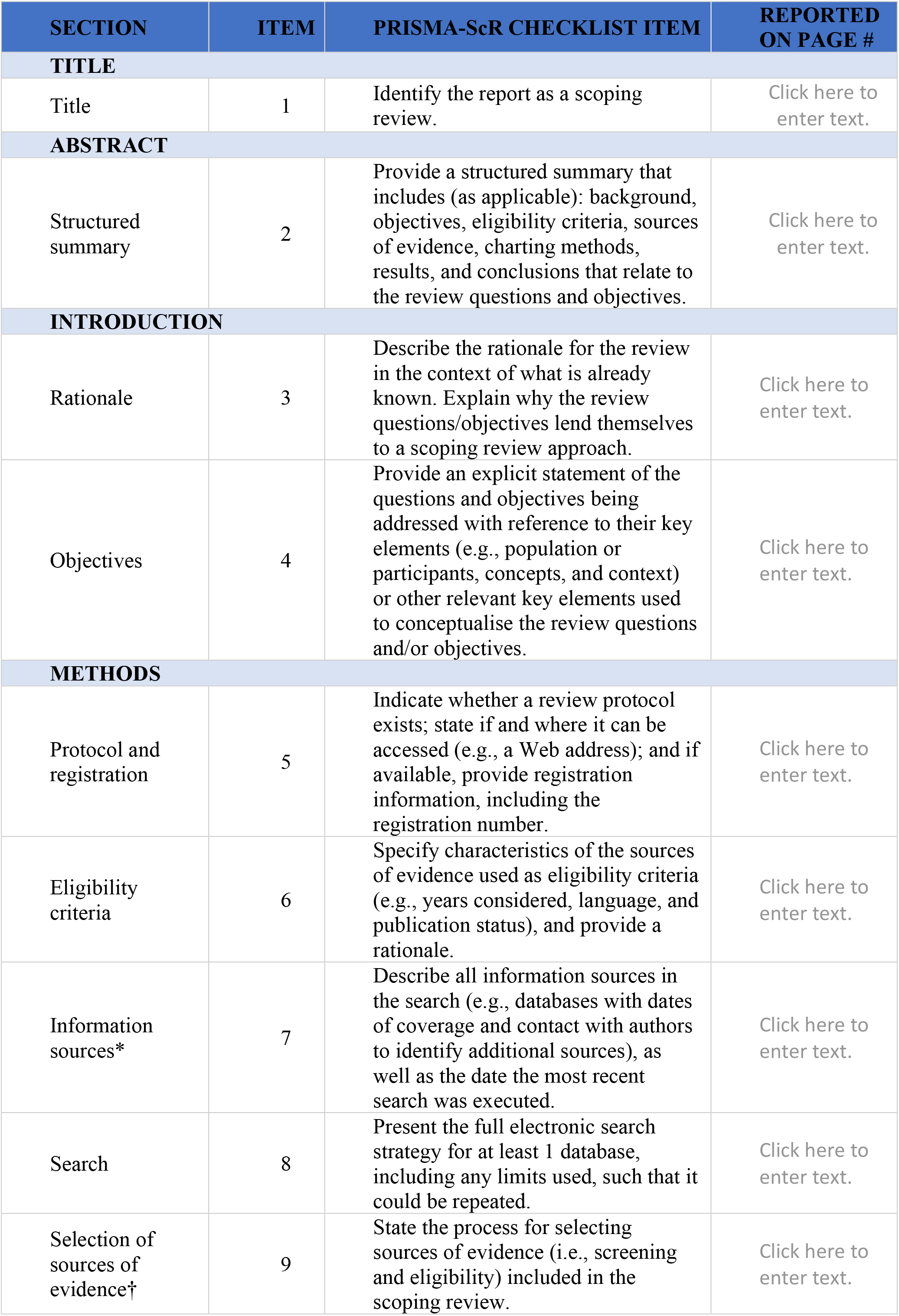
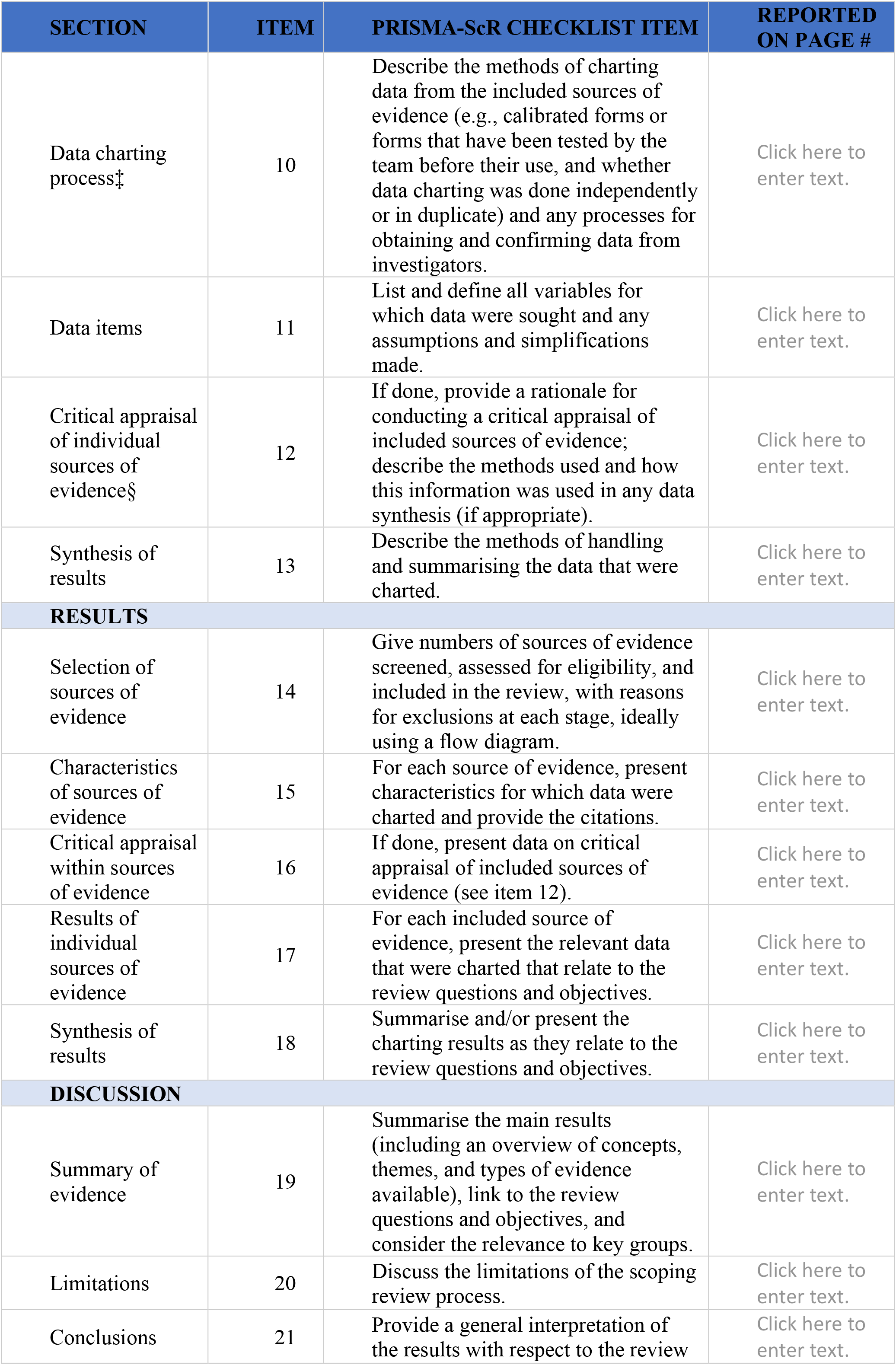
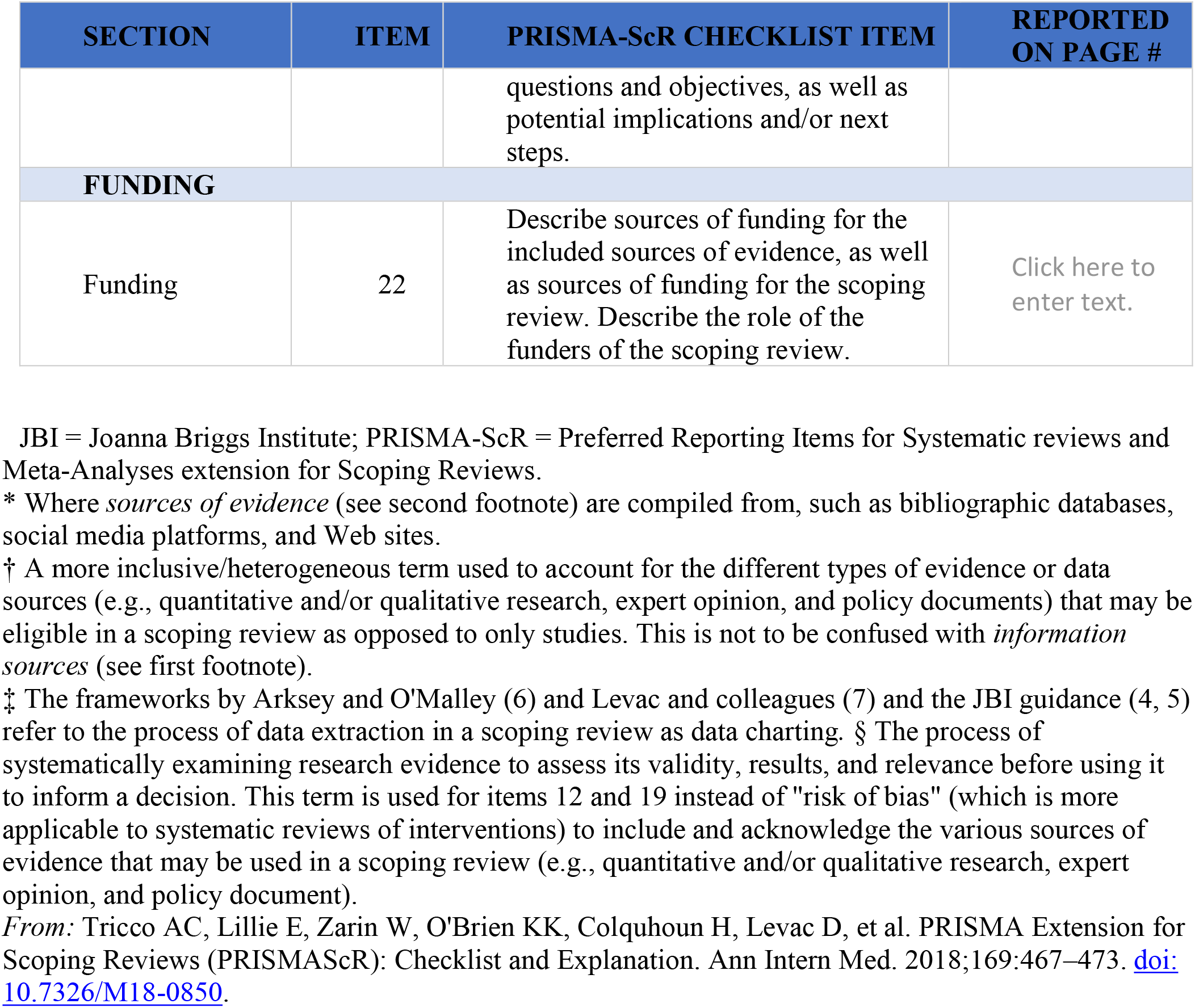
PRISMA-ScR Checklist.

**Supplementary Figure 1:**
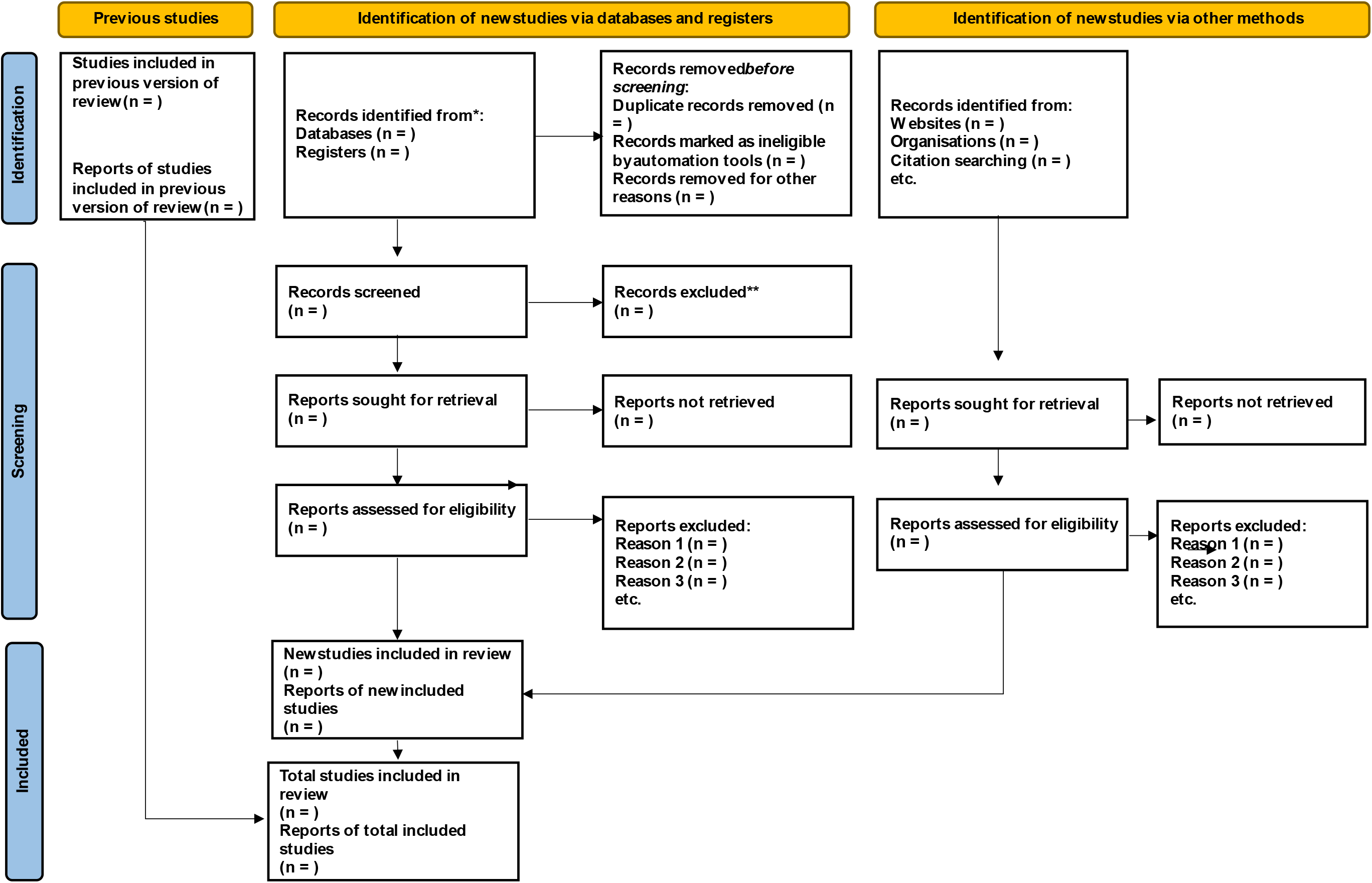
PRISMA 2020 flow diagram for the study selection process.

## Footnotes

1 Qualitive studies will be excluded because the scoping review will focus on consolidating evidence on estimations, measurements and distributions of the concept of the research question.

## Notes

### Competing Interest Statement

The authors have declared no competing interest.

### Funding Statement

This study did not receive any funding

