## Supplemental Data 1 for "Molecular Epidemiology and Histopathological Classifications of Lung Cancer in Africa: A Scoping Review Protocol"

**APPENDIX: Supplementary Files**

**Supplementary Table 1: Pilot search in PubMed electronic database**

| **Date** | **Database** | **Keywords** | **Search results** |
|---|---|---|---|
| 7/06/2022 | PubMed | ("molecular epidemiology"[MeSH Terms] OR ("molecular"[All Fields] AND "epidemiology"[All Fields]) OR "molecular epidemiology"[All Fields]) AND histopathological[All Fields] AND ("classification"[Subheading] OR "classification"[All Fields] OR "classification"[MeSH Terms]) AND ("lung neoplasms"[MeSH Terms] OR ("lung"[All Fields] AND "neoplasms"[All Fields]) OR "lung neoplasms"[All Fields] OR ("lung"[All Fields] AND "cancer"[All Fields]) OR "lung cancer"[All Fields]) AND ("africa"[MeSH Terms] OR "africa"[All Fields]) | 718 |

**Supplementary Table 2:**

| **Table: Data extraction form** |
|---|
| Author and date of publication |
| Study title |
| Aim(s) of the study or research questions |
| Study design |
| Study setting (urban/rural) |
| Population<br>- Average age<br>- Gender<br>- Sample size<br>- percentage of women<br>- percentage of men |
| Methodology/intervention |
| Type of Intervention and outcomes |
| Most relevant finding |
| Most significant finding |
| Other key findings |
| Study limitations and implications |
| Interpretations and conclusions from the authors |
| Comment/s |

**Supplementary Table 3: PRISMA-ScR Checklist**

| SECTION | ITEM | PRISMA-ScR CHECKLIST ITEM | REPORTED ON PAGE # |
|---|---|---|---|
| **TITLE** | | | |
| Title | 1 | Identify the report as a scoping review. | Click here to enter text. |
| **ABSTRACT** | | | |
| Structured summary | 2 | Provide a structured summary that includes (as applicable): background, objectives, eligibility criteria, sources of evidence, charting methods, results, and conclusions that relate to the review questions and objectives. | Click here to enter text. |
| **INTRODUCTION** | | | |
| Rationale | 3 | Describe the rationale for the review in the context of what is already known. Explain why the review questions/objectives lend themselves to a scoping review approach. | Click here to enter text. |
| Objectives | 4 | Provide an explicit statement of the questions and objectives being addressed with reference to their key elements (e.g., population or participants, concepts, and context) or other relevant key elements used to conceptualise the review questions and/or objectives. | Click here to enter text. |
| **METHODS** | | | |
| Protocol and registration | 5 | Indicate whether a review protocol exists; state if and where it can be accessed (e.g., a Web address); and if available, provide registration information, including the registration number. | Click here to enter text. |
| Eligibility criteria | 6 | Specify characteristics of the sources of evidence used as eligibility criteria (e.g., years considered, language, and publication status), and provide a rationale. | Click here to enter text. |
| Information sources* | 7 | Describe all information sources in the search (e.g., databases with dates of coverage and contact with authors to identify additional sources), as well as the date the most recent search was executed. | Click here to enter text. |
| Search | 8 | Present the full electronic search strategy for at least 1 database, including any limits used, such that it could be repeated. | Click here to enter text. |
| Selection of sources of evidence† | 9 | State the process for selecting sources of evidence (i.e., screening and eligibility) included in the scoping review. | Click here to enter text. |

| SECTION | ITEM | PRISMA-ScR CHECKLIST ITEM | REPORTED ON PAGE # |
| --- | --- | --- | --- |
| Data charting process‡ | 10 | Describe the methods of charting data from the included sources of evidence (e.g., calibrated forms or forms that have been tested by the team before their use, and whether data charting was done independently or in duplicate) and any processes for obtaining and confirming data from investigators. | Click here to enter text. |
| Data items | 11 | List and define all variables for which data were sought and any assumptions and simplifications made. | Click here to enter text. |
| Critical appraisal of individual sources of evidence§ | 12 | If done, provide a rationale for conducting a critical appraisal of included sources of evidence; describe the methods used and how this information was used in any data synthesis (if appropriate). | Click here to enter text. |
| Synthesis of results | 13 | Describe the methods of handling and summarising the data that were charted. | Click here to enter text. |
| **RESULTS** | | | |
| Selection of sources of evidence | 14 | Give numbers of sources of evidence screened, assessed for eligibility, and included in the review, with reasons for exclusions at each stage, ideally using a flow diagram. | Click here to enter text. |
| Characteristics of sources of evidence | 15 | For each source of evidence, present characteristics for which data were charted and provide the citations. | Click here to enter text. |
| Critical appraisal within sources of evidence | 16 | If done, present data on critical appraisal of included sources of evidence (see item 12). | Click here to enter text. |
| Results of individual sources of evidence | 17 | For each included source of evidence, present the relevant data that were charted that relate to the review questions and objectives. | Click here to enter text. |
| Synthesis of results | 18 | Summarise and/or present the charting results as they relate to the review questions and objectives. | Click here to enter text. |
| **DISCUSSION** | | | |
| Summary of evidence | 19 | Summarise the main results (including an overview of concepts, themes, and types of evidence available), link to the review questions and objectives, and consider the relevance to key groups. | Click here to enter text. |
| Limitations | 20 | Discuss the limitations of the scoping review process. | Click here to enter text. |
| Conclusions | 21 | Provide a general interpretation of the results with respect to the review | Click here to enter text. |

| SECTION | ITEM | PRISMA-ScR CHECKLIST ITEM | REPORTED ON PAGE # |
|---|---|---|---|
| | | questions and objectives, as well as potential implications and/or next steps. | |
| **FUNDING** | | | |
| Funding | 22 | Describe sources of funding for the included sources of evidence, as well as sources of funding for the scoping review. Describe the role of the funders of the scoping review. | Click here to enter text. |

JBI = Joanna Briggs Institute; PRISMA-ScR = Preferred Reporting Items for Systematic reviews and Meta-Analyses extension for Scoping Reviews.
* Where *sources of evidence* (see second footnote) are compiled from, such as bibliographic databases, social media platforms, and Web sites.
† A more inclusive/heterogeneous term used to account for the different types of evidence or data sources (e.g., quantitative and/or qualitative research, expert opinion, and policy documents) that may be eligible in a scoping review as opposed to only studies. This is not to be confused with *information sources* (see first footnote).
‡ The frameworks by Arksey and O'Malley (6) and Levac and colleagues (7) and the JBI guidance (4, 5) refer to the process of data extraction in a scoping review as data charting. § The process of systematically examining research evidence to assess its validity, results, and relevance before using it to inform a decision. This term is used for items 12 and 19 instead of "risk of bias" (which is more applicable to systematic reviews of interventions) to include and acknowledge the various sources of evidence that may be used in a scoping review (e.g., quantitative and/or qualitative research, expert opinion, and policy document).
*From:* Tricco AC, Lillie E, Zarin W, O'Brien KK, Colquhoun H, Levac D, et al. PRISMA Extension for Scoping Reviews (PRISMAScR): Checklist and Explanation. Ann Intern Med. 2018;169:467–473. doi: 10.7326/M18-0850.

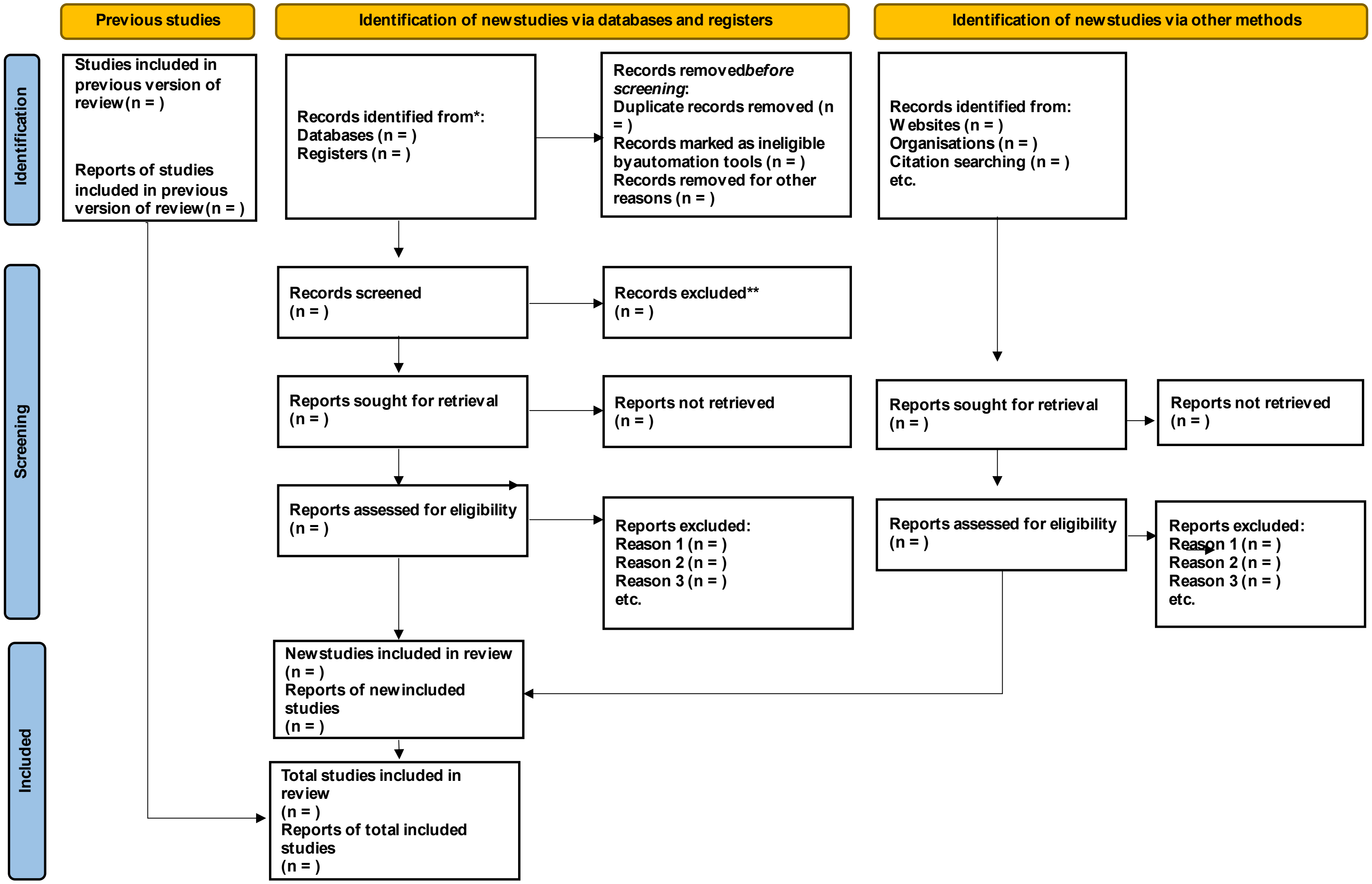


**Supplementary Figure 1:** PRISMA 2020 flow diagram for the study selection process
